# Effects of Anticoagulants and Corticosteroids therapy in patients affected by severe COVID-19 Pneumonia

**DOI:** 10.1101/2020.06.22.20134957

**Authors:** Khalid Ghalilah, Abdul Momin Sabir, Irshad Ali Alvi, Malak Alharbi, Abdulrahman Basabrain, Mahmooud Aljundi, Ghazi Almohammadi, Zainab Almuairfi, Raed Alharbi

## Abstract

**Background:** In the absence of a standard of treatment for COVID-19, the combined use of anti-inflammatory (corticosteroids and Enoxaparin) and antiviral drugs may be more effective than using either modality alone in the treatment of COVID-19.

**Methods:** Patients hospitalized between April 10th, 2020, through May 10th, 2020, who had confirmed COVID-19 infection with clinical or radiographic evidence of pneumonia, in which 65 patients have moderate COVID-19 pneumonia, and 63 patients have severe COVID-19 pneumonia. All patients received early combination therapy of anti-inflammatory (corticosteroids and Enoxaparin) and antiviral drugs. They assessed for type and duration of treatment, and days need to wean from oxygen therapy, length of stay, virus clearance time, and complication or adverse events. All patients had more than 28 days follow up after discharge from the hospital.

**Results:** Moderate COVID-19 pneumonia group were 65 patients who received Enoxaparin, antiviral drugs, empirical antibiotics for pneumonia, and standard treatment for comorbidity. Male patients were 50 (76.9 %) and female patients were 15 (23.1 %). 34 (52.3 %) patients have comorbidity, 25 (38.5%) patients have Diabetes Mellitus and 2 (3.1 %) pregnant ladies. 19 (29.2 %) patients were on low flow oxygen therapy, 3L oxygen or less to maintain oxygen saturation more than 92%. All patients discharged home with no major or minor bleeding complications or significant complications. Severe COVID-19 pneumonia group were 63 patients who received methylprednisolone, enoxaparin, antiviral drugs, empirical antibiotics for pneumonia, and standard treatment for comorbidity. Male patients were 55 (87.3 %) and female patients were 8 (12.7 %). 37 (58.7 %) patients have comorbidity, and 24 (38.1%) patients have Diabetes Mellitus. 32 (50.8 %) patients were on low flow oxygen therapy, 4-9L oxygen, and 31 (49.2 %) patients were on low flow oxygen therapy, 10L oxygen or more, including 12 patients on a non-rebreathing mask. Patients received methylprednisolone were 37 (58.7 %) for 3 days, 16 (25.4 %) for 5 days and 10 (15.9 %) for more than 5 days. Sixty-two patients discharged home with one patient had a long stay, and the other two transferred to ICU. One long-stay patient transferred to ICU on low flow oxygen therapy.

**Conclusion:** Early use of a combined anti-inflammatory (corticosteroids and Enoxaparin) and antiviral drugs treatment in patients with moderate to severe COVID-19 pneumonia prevent complications of the disease and improve clinical outcomes

## Introduction

The Novel Coronavirus 2019 was first reported on in Wuhan, China, in late December 2019. The outbreak was declared a public health emergency of international concern in January 2020, and on March 11^th^, 2020, the outbreak was declared a global pandemic1. The virus has been named severe acute respiratory syndrome coronavirus-2 (SARS-CoV-2), and the disease it causes has become known as coronavirus disease 2019 (COVID-19).^1^

An estimated 20% of COVID-19 cases are sick enough to require hospitalization, with a subset of 5% of patients requiring intensive care. Although most reported COVID-19 cases in China were mild (81%).^2^ Among hospitalized patients with COVID-19, about 10-20% of patients are admitted to ICU, 3-10% require intubation, and 2-5% die.^3^ As of April 30th, the Case Fatality Rate prediction interval is 0.82-9.64%.^4^

To date, there is no specific antiviral drug or vaccine for COVID-19. All the treatment options come from experience treating SARS, MERS, or some other new influenza virus previously. Treatments are mainly symptomatic and respiratory support. There is no current evidence from RCTs to recommend any specific treatment for patients with suspected or confirmed COVID-19.

No studies were found explicitly examining the role of steroids for the treatment of the COVID-19 pneumonia. Corticosteroids were widely used in China to prevent the development of ARDS in patients with COVID-19 pneumonia. Among patients who have been admitted to the hospital with COVID-19 pneumonia, the IDSA guideline panel suggests against the use of corticosteroids.^5^ Surviving Sepsis Campaign guideline on managing critically ill adults with COVID-19 supports using corticosteroids in mechanically ventilated patients with COVID-19 and acute respiratory distress syndrome (but not those with respiratory failure in the absence of that syndrome) and patients with COVID-19 and refractory shock; short-course, low-dose regimens are preferred.^6^

In the absence of a standard of treatment for COVID-19, the combined use of anti-inflammatory (corticosteroids and Enoxaparin) and antiviral drugs may be more effective than using either modality alone in the treatment of COVID-19. This study evaluates the effectiveness of a combination treatment of anti-inflammatory (corticosteroids and Enoxaparin) and antiviral drugs to treat severe COVID-19 pneumonia.

## Methods

### Study setting, patients and Design

This study was done at Al Madinah Al Munawarah Hospital in Al Madinah, Saudi Arabia, from April 10^th^ to June 10^th^, 2020. Patients were received and admitted through the Emergency Department. Cases were positive for SARS-CoV-2 by RT-qPCR with findings of pneumonia clinically or radiologically on a chest x-ray or CT scan. All patients were followed by the infectious diseases team from admission to discharge, and more than 28 days post-discharge follow up for complications or readmission. All patients signed informed consent for receiving medical acts and care.

All patients hospitalized between April 10^th^, 2020 through May ^10^th, 2020, were eligible for inclusion if they were 18 years of age or older, had confirmed COVID-19 infection with clinical or radiographic evidence of pneumonia. Patients were excluded if they were immunocompromised with cancer, chemotherapy, transplant, or end-stage renal disease on renal replacement therapy.

### Definitions

A confirmed case of COVID-19 was defined as a patient that had a positive reverse-transcriptase– polymerase-chain-reaction (RT-PCR) assay for SARS-CoV-2 in a nasopharyngeal sample tested by Al-Madinah region central lab.

Severe COVID-19 pneumonia defined as confirmed COVID-19 infection, with clinical or radiographic evidence of pneumonia, and oxygen saturation less than 90 with pulse oximetry.

Moderate COVID-19 pneumonia defined as confirmed COVID-19 infection, with clinical or radiographic evidence of pneumonia, and oxygen saturation more than 90 with pulse oximetry.

### Treatment

1. Intravenous methylprednisolone: only for severe COVID-19 pneumonia; if the patient needs more than 10L oxygen therapy, methylprednisolone is 40mg four times daily for 3 to 5 days, if the patient does not improve, continue until his oxygen saturation becomes more than 90% on ambient air, then methylprednisolone is tapering to once daily and to be stopped if oxygen saturation more than 93% on ambient air. If the patient needs less than 10L oxygen therapy, methylprednisolone is 40mg three times daily for 3 to 5 days if the patient does not improve, continue until his oxygen saturation becomes more than 90% on ambient air, then methylprednisolone is tapering to once daily and to be stopped if oxygen saturation more than 93% on ambient air.
2. Enoxaparin treatment according to D-Dimer, patient weight, and severity of pneumonia.

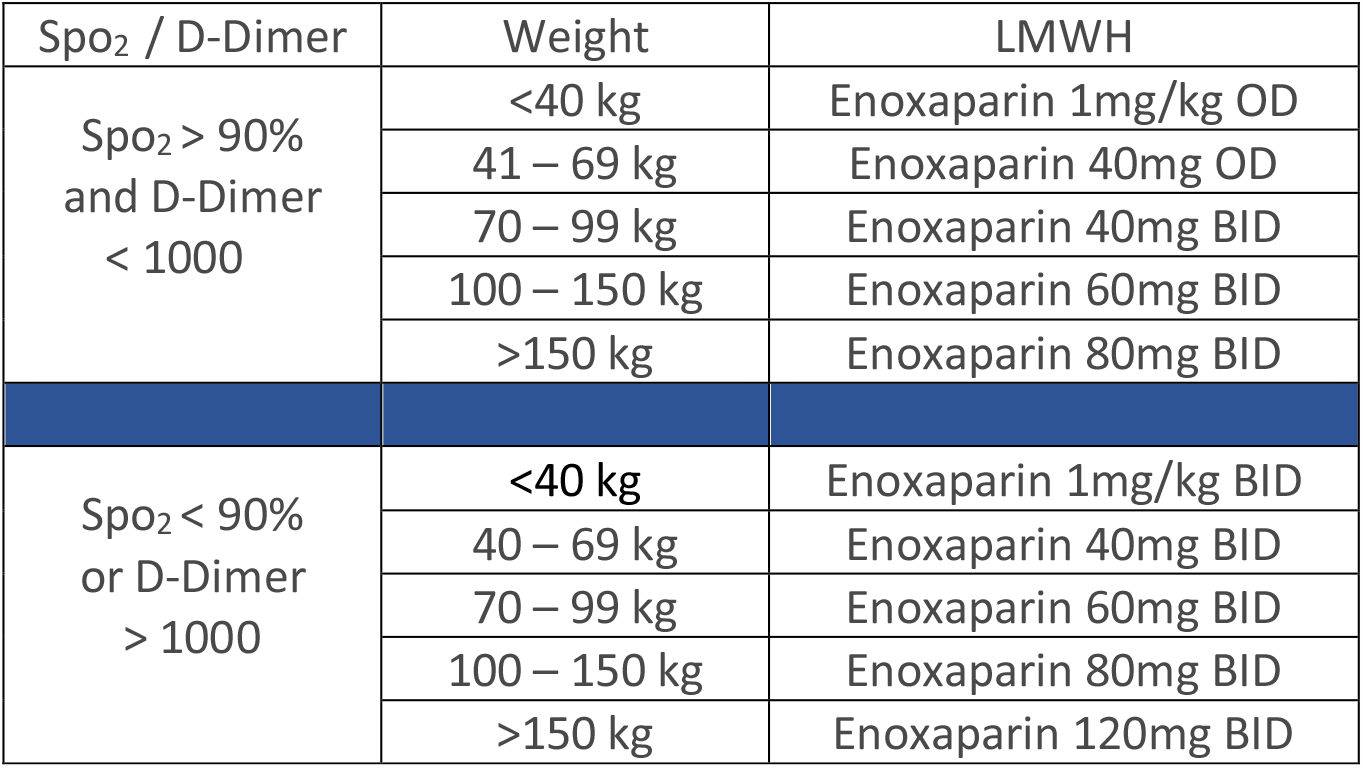 Duration of Enoxaparin treatment: Moderate COVID-19 pneumonia is three to five days; if the patient does not improve, continue enoxaparin until oxygen saturation becomes more than 93% on ambient air, then change enoxaparin dose to 40mg once daily until home discharge. Enoxaparin duration for severe COVID-19 pneumonia is three to five days, if the patient doesn’t improve, continue enoxaparin until oxygen saturation becomes more than 90% on ambient air, then decrease the enoxaparin according to his weight, continue enoxaparin until oxygen saturation becomes more than 93% on ambient air, then change enoxaparin dose to 40mg once daily until home discharge.
3. Antiviral drugs were hydroxychloroquine 400 mg every 12 hours for one day, followed by 200 mg twice daily for 5 – 7 days or Lopinavir/Ritonavir 400/100 mg twice daily for ten days.
4. Empirical antibiotics for pneumonia.
5. Other treatments: standard treatment for comorbidity.

### Study Assessments

All patients assessed for type and duration of treatment, days need to wean from oxygen therapy, length of stay, virus clearance time, and complication or adverse events. All patients had 28 days follow up or more after discharge from the hospital by mobile phone or emergency department visit for hospital readmission or complication.

### Outcome definitions

1. ICU transfer after admission in the medical ward due to respiratory failure or any reason.
2. Patient discharged home.
3. Long stay mor than 21 days.
4. Death due to any cause.

### Statistical analysis

Data were analyzed using IBM SPSS for Windows. Statistical analyses of demographics, clinical, laboratory, treatment, length of stay, virus clearance time, and outcome descriptive data are tabulated. Descriptive statistics such as means and standard deviation mean (±SD) were used for quantitative variables, absolute and relative frequency (%) for qualitative variables.

## Results

### Characteristics of the Patients

Between April 10^th^ to May 10^th^, 2020, A total of 128 patients with COVID-19 were enrolled, in which 65 patients have moderate COVID-19 pneumonia, and 63 patients have severe COVID-19 pneumonia. Table 1 shows the baseline demographic and clinical characteristics of both groups.

**Table (1).** Baseline Demographics and Clinical Characteristics of Study Patients

| Characteristics | Covid-19 Pneumonia without hypoxemia $\geq 90\%$<br>(including patients need 3L oxygen or less to maintain oxygen saturation more than 92%) | Covid-19 Pneumonia with Hypoxemia $< 90\%$<br>(All patients need more than 3L oxygen therapy to maintain oxygen saturation more than 92%) |
| --- | --- | --- |
| **Age – no. (%)** |  |  |
| 18-40 | 14 (21.5 %) | 15 (23.8 %) |
| 41-60 | 38 (58.5 %) | 39 (61.9 %) |
| 60-80 | 13 (20.0 %) | 9 (14.3 %) |
| **Sex – no. (%)** |  |  |
| Male | 50 (76.9 %) | 55 (87.3 %) |
| Female | 15 (23.1 %) | 8 (12.7 %) |
| **Comorbidity or Coexisting conditions – no. (%)** |  |  |
| NO | 31 (47.7 %) | 26 (41.3 %) |
| YES | 34 (52.3 %) | 37 (58.7 %) |
| DM | 15 (23.1 %) | 15 (23.8 %) |
| HTN | 4 (6.2 %) | 8 (12.7 %) |
| DM+HTN | 4 (6.2 %) | 6 (9.5 %) |
| DM+IHD | 1 (1.5 %) | 1 (1.6 %) |
| DM+HTN+ICM | 2 (3.1 %) | 1 (1.6 %) |
| DM+HTN+CKD | 1 (1.5 %) | 0 |
| DM+HTN+Stroke | 1 (1.5 %) | 0 |
| HTN+CKD | 1 (1.5 %) | 0 |
| HTN+IHD | 0 | 1 (1.6 %) |
| DM+COPD/B. Asthma | 1 (1.5 %) | 1 (1.6 %) |
| HTN+COPD/B. Asthma | 1 (1.5 %) | 2 (3.2 %) |
| Morbid Obesity | 1 (1.5 %) | 1 (1.6 %) |
| Sickle cell Anemia | 0 | 1 (1.6 %) |
| Pregnancy | 2 (3.1 %) | 0 |
| **D-dimer Level – no. (%)** |  |  |
| < 1000 | 47 (72.3%) | 38 (60.3 %) |
| 1000 – 3000 | 15 (23.1 %) | 21 (33.3 %) |
| > 3000 | 3 (4.6 %) | 4 (6.3 %) |
| **Oxygen Therapy at Admission – no. (%)** |  |  |
| Ambient Air | 46 (70.8 %) | 0 |
| Low Flow Oxygen<br>3L oxygen or less | 19 (29.2 %) | 0 |
| Low Flow Oxygen<br>4L – 9 L oxygen | 0 | 32 (50.8 %) |
| Low Flow Oxygen<br>10L oxygen or more | 0 | 31 (49.2 %) <sup>a</sup> |
| **Anti-inflammatory and Antiviral Treatment – no. (%)** |  |  |
| Anticoagulant | 9 (13.8%) | 0 |
| Anticoagulant + Hydroxychloroquine | 56 (86.2 %) | 0 |
| Corticosteroids + Anticoagulant + Hydroxychloroquine | 0 | 59 (93.7 %) |
| Corticosteroids + Anticoagulant + Lopinavir | 0 | 4 (6.3 %) |
| **Days of Corticosteroids Treatment – no. (%)** |  |  |
| 3 days | 0 | 37 (58.7 %) |
| 5 days | 0 | 16 (25.4 %) |
| More than 5 days | 0 | 10 (15.9 %) |
| **Empirical Antibiotics – no. (%)** |  |  |
| Ceftriaxone + Azithromycin or Levofloxacin | 48 (73.8 %) | 3 (4.8 %) |
| Cefepime or Meropenem or PIP-TAZO + Azithromycin or Levofloxacin | 14 (21.5 %) | 42 (66.6 %) |
| Cefepime or Meropenem or PIP-TAZO + Azithromycin or Levofloxacin + Vancomycin or Linezolid | 3 (4.6 %) | 18 (28.6 %) |
| **Azithromycin + Hydroxychloroquine – no. (%)** |  |  |
| YES | 42 (64.6 %) | 50 (79.4 %) |
| NO | 23 (35.4 %) | 13 (20.6 %) |
| **Days need to wean from Oxygen Therapy – no. (%)** |  |  |
| 2-3 | 6 (9.2 %) | 15 (23.8 %) |
| 4-5 | 9 (13.8 %) | 13 (20.6 %) |
| 6-7 | 1 (1.5 %) | 7 (11.1 %) |
| 8-10 | 1 (1.5 %) | 15 (23.8 %) |
| 11-14 | 1 (1.5 %) | 9 (14.3 %) |
| more than 14 days | 1 (1.5 %) | 4 (6.3 %) |
| **Outcome – no. (%)** |  |  |
| No ICU +Discharge Home | 65 (100 %) | 59 (93.6 %) |
| Yes ICU +Discharge Home | 0 | 2 (3.2 %) |
| No ICU + Long Stay | 0 | 1 (1.6 %) |
| Yes ICU + Long Stay | 0 | 1 (1.6 %) |
| Death | 0 | 0 |
| **Mean of length of stay in days (Maximum- Minimum)** |  |  |
|  | 6.7 days (18 – 3) | 9.7 days (30 – 3) |
| **Mean of virus clearance time in days (Maximum- Minimum)** |  |  |
|  | 14.3 days (24 – 10) | 14.3 (27 – 6) |
| **Complication or Adverse Events – no. (%)** |  |  |
| NO | 54 (83 %) | 39 (62 %) |
| YES | 11 (17 %) | 24 (38 %) |
| Major Bleeding (GIT bleeding or ICH) |  |  |
| Minor Bleeding |  |  |
| Septic shock |  |  |
| Bacteremia | 1 (1.5 %) |  |
| Acute Respiratory Failure |  |  |
| Pulmonary Embolism |  |  |
| Lung Abscess or Empyema |  |  |
| Acute Coronary Syndrome |  |  |
| Arrhythmia | 1 (1.5 %) |  |
| Acute Kidney Injury | 1 (1.5 %) | 2 (3.2 %) |
| Nausea or Vomiting | 2 (3.1 %) | 2 (3.2 %) |
| Gastritis / Duodenitis |  |  |
| Diarrhea | 4 (6.2 %) | 2 (3.2 %) |
| Constipation |  |  |
| Elevated ALT or AST | 2 (3.1 %) |  |
| Hyperglycemia |  | 15 (23.8 %) |
| Hypokalemia |  |  |
| Hyperglycemia + Hypokalemia |  | 3 (4.8 %) |
| Hypernatremia |  |  |
| Anemia |  |  |
| Coagulopathy |  |  |
| Hypertension |  |  |

<sup>a</sup> including 12 patients on non-rebreathing mask

**Table (2).**
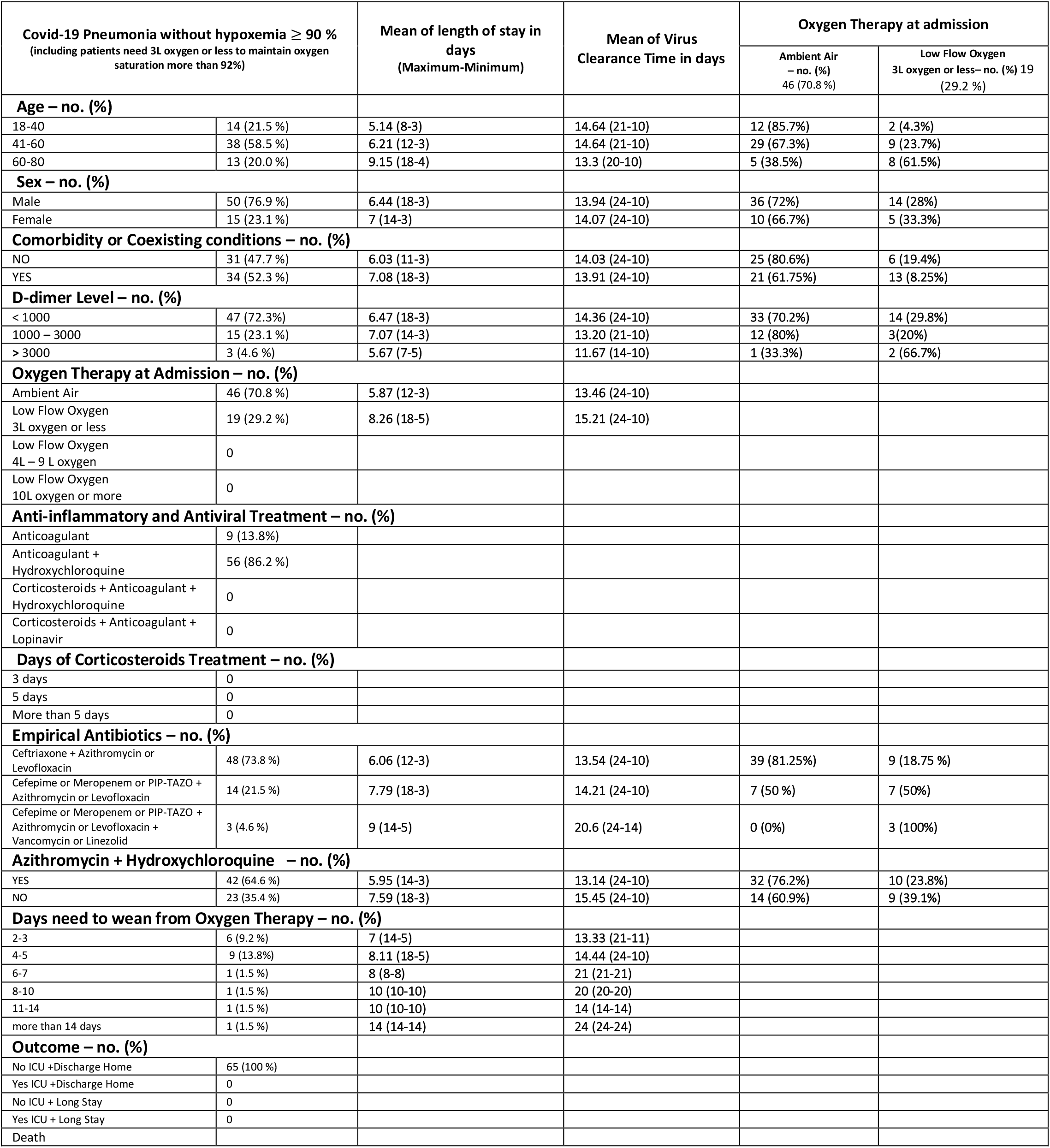
Covid 19 Pneumonia without hypoxemia and characteristics of Mean of length of stay, Mean of virus clearance and Oxygen therapy at admission

**Table (3).** Covid 19 Pneumonia without hypoxemia and characteristics of Empirical Antibiotics

| Covid-19 Pneumonia without hypoxemia $\geq 90\%$<br>(including patients need 3L oxygen or less to maintain oxygen saturation more than 92%) | | Empirical Antibiotics | | |
| --- | --- | --- | --- | --- |
|  |  | Ceftriaxone + Azithromycin or Levofloxacin<br>No. (%) / 48 (73.8 %) | Cefepime or Meropenem or PIP-TAZO + Azithromycin or Levofloxacin<br>No. (%) / 14 (21.5 %) | Cefepime or Meropenem or PIP-TAZO + Azithromycin or Levofloxacin + Vancomycin or Linezolid<br>No. (%) / 3 (4.6 %) |
| **Age – no. (%)** |  |  |  |  |
| 18-40 | 14 (21.5 %) | 11 (78.6%) | 2 (14.3%) | 1 (7.1%) |
| 41-60 | 38 (58.5 %) | 29 (76.3%) | 9 (23.7%) | 0 (0%) |
| 60-80 | 13 (20.0 %) | 8 (61.5%) | 3 (23.1%) | 2 (15.4%) |
| **Sex – no. (%)** |  |  |  |  |
| Male | 50 (76.9 %) | 35 (70%) | 12 (24%) | 3 (6%) |
| Female | 15 (23.1 %) | 13 (68.7%) | 2 (13.3%) | 0 (0%) |
| **Comorbidity or Coexisting conditions – no. (%)** |  |  |  |  |
| NO | 31 (47.7 %) | 26 (83.9%) | 4 (12.9%) | 1 (3.2%) |
| YES | 34 (52.3 %) | 22 (64.7%) | 10 (29.4%) | 2 (5.9%) |
| **D-dimer Level – no. (%)** |  |  |  |  |
| < 1000 | 47 (72.3%) | 35 (74.5%) | 9 (19.1%) | 3 (6.4%) |
| 1000 – 3000 | 15 (23.1 %) | 11 (73.3%) | 4 (26.7%) | 0 (0%) |
| > 3000 | 3 (4.6 %) | 2 (66.7%) | 1 (33.3%) | 0 (0%) |
| **Oxygen Therapy at Admission – no. (%)** |  |  |  |  |
| Ambient Air | 46 (70.8 %) | 39 (84.8%) | 7 (15.2%) | 0 |
| Low Flow Oxygen<br>3L oxygen or less | 19 (29.2 %) | 9 (47.4%) | 7 (36.8%) | 3 (15.8%) |
| Low Flow Oxygen<br>4L – 9 L oxygen | 0 |  |  |  |
| Low Flow Oxygen<br>10L oxygen or more | 0 |  |  |  |
| **Anti-inflammatory and Antiviral Treatment – no. (%)** |  |  |  |  |
| Anticoagulant | 9 (13.8%) |  |  |  |
| Anticoagulant + Hydroxychloroquine | 56 (86.2 %) |  |  |  |
| Corticosteroids + Anticoagulant + Hydroxychloroquine | 0 |  |  |  |
| Corticosteroids + Anticoagulant + Lopinavir | 0 |  |  |  |
| **Days of Corticosteroids Treatment – no. (%)** |  |  |  |  |
| 3 days | 0 |  |  |  |
| 5 days | 0 |  |  |  |
| More than 5 days | 0 |  |  |  |
| **Empirical Antibiotics – no. (%)** |  |  |  |  |
| Ceftriaxone + Azithromycin or Levofloxacin | 48 (73.8 %) |  |  |  |
| Cefepime or Meropenem or PIP-TAZO + Azithromycin or Levofloxacin | 14 (21.5 %) |  |  |  |
| Cefepime or Meropenem or PIP-TAZO + Azithromycin or Levofloxacin + Vancomycin or Linezolid | 3 (4.6 %) |  |  |  |
| **Azithromycin + Hydroxychloroquine – no. (%)** |  |  |  |  |
| YES | 42 (64.6 %) | 36 (85.7%) | 6 (14.3%) | 0 (0%) |
| NO | 23 (35.4 %) | 12 (52.2%) | 8 (34.8%) | 3 (13%) |
| **Days need to wean from Oxygen Therapy – no. (%)** |  |  |  |  |
| 2-3 | 6 (9.2 %) | 2 (33.3%) | 4 (66.7%) | 0 (0%) |
| 4-5 | 9 (13.8%) | 6 (66.7%) | 2 (22.2%) | 1 (11.1%) |
| 6-7 | 1 (1.5 %) | 0 (0%) | 0 (0%) | 1 (100%) |
| 8-10 | 1 (1.5 %) | 0 (0%) | 1 (100%) | 0 (0%) |
| 11-14 | 1 (1.5 %) | 1 (100%) | 0 (0%) | 0 (0%) |
| more than 14 days | 1 (1.5 %) | 0 (0%) | 0 (0%) | 1 (100%) |
| **Outcome – no. (%)** |  |  |  |  |
| No ICU +Discharge Home | 65 (100 %) |  |  |  |
| Yes ICU +Discharge Home | 0 |  |  |  |
| No ICU + Long Stay | 0 |  |  |  |
| Yes ICU + Long Stay | 0 |  |  |  |
| Death | 0 |  |  |  |

**Table (4).** Severe Covid 19 Pneumonia with hypoxemia and characteristics of mean of length of stay, Mean of virus clearance and Oxygen therapy at admission

| Covid-19 Pneumonia with Hypoxemia < 90 %<br>(All patients need more than 3L oxygen therapy to maintain oxygen saturation more than 92%) |  | Mean of length of stay in days<br>(Maximum-Minimum) | Mean of Virus Clearance Time in days | Oxygen Therapy at admission |  |
| --- | --- | --- | --- | --- | --- |
|  |  |  |  | Low Flow Oxygen<br>4L – 9 L oxygen – no. (%)<br>32 (50.8 %) | Low Flow Oxygen<br>10L oxygen or more – no. (%)<br>31 (49.2 %) |
| **Age – no. (%)** |  |  |  |  |  |
| 18-40 | 15 (23.8 %) | 9 (16-5) | 113.27 (20-5) | 5 (33.3%) | 10 (66.7%) |
| 41-60 | 39 (61.9 %) | 10.62 (30-5) | 15.03 (27-6) | 23 (59%) | 16 (41%) |
| 60-80 | 9 (14.3 %) | 11 (18-10) | 13.78 (18-10) | 4 (44.4%) | 5 (55.6%) |
| **Sex – no. (%)** |  |  |  |  |  |
| Male | 55 (87.3 %) | 7 (14-3) | 10.47 (24-10) | 26 (47.3%) | 29 (52.7%) |
| Female | 8 (12.7 %) | 9 (13-5) | 13.13 (21-5) | 6 (75%) | 2 (25%) |
| **Comorbidity or Coexisting conditions – no. (%)** |  |  |  |  |  |
| NO | 26 (41.3 %) | 8.85 (17-5) | 14.23 (27-6) | 16 (61.5%) | 10 (38.5%) |
| YES | 37 (58.7 %) | 11.30 (30-6) | 14.57 (24-5) | 15 (40.5%) | 22 (59.5%) |
| **D-dimer Level – no. (%)** |  |  |  |  |  |
| < 1000 | 38 (60.3 %) | 11.32 (30-5) | 14.92 (24-8) | 16 (42.1%) | 22 (57.9%) |
| 1000 – 3000 | 21 (33.3 %) | 8.38 (17-5) | 13.48 (27-5) | 14 (66.7%) | 7 (33.3%) |
| > 3000 | 4 (6.3 %) | 10.50 (14-5) | 14.75 (18-12) | 2 (50%) | 2 (50%) |
| **Oxygen Therapy at Admission – no (%)** |  |  |  |  |  |
| Ambient Air | 0 |  |  |  |  |
| Low Flow Oxygen<br>3L oxygen or less | 0 |  |  |  |  |
| Low Flow Oxygen<br>4L – 9 L oxygen | 32 (50.8 %) | 8.03 (12-5) | 13.8 (27-5) |  |  |
| Low Flow Oxygen<br>10L oxygen or more | 31 (49.2 %)<sup>a</sup> | 12.47 (30-5) | 15.03 (24-8) |  |  |
| **Anti-inflammatory and Antiviral Treatment – no. (%)** |  |  |  |  |  |
| Anticoagulant | 0 |  |  |  |  |
| Anticoagulant + Hydroxychloroquine | 0 |  |  |  |  |
| Corticosteroids + Anticoagulant + Hydroxychloroquine | 59 (93.7 %) |  |  |  |  |
| Corticosteroids + Anticoagulant + Lopinavir | 4 (6.3 %) |  |  |  |  |
| **Days of Corticosteroids Treatment – no. (%)** |  |  |  |  |  |
| 3 days | 37 (58.7 %) | 7.96 (16-5) | 13.78 (27-5) | 25 (67.6%) | 12 (32.4%) |
| 5 days | 16 (25.4 %) | 12.25 (30-7) | 14.81 (24-8) | 6 (37.5%) | 10 (62.5%) |
| More than 5 days | 10 (15.9 %) | 15.7 (30-6) | 16.20 (24-8) | 0 (0%) | 10 (100%) |
| **Empirical Antibiotics – no. (%)** |  |  |  |  |  |
| Ceftriaxone + Azithromycin or Levofloxacin | 3 (4.8 %) | 7 (10-5) | 13.67 (19-10) | 3 (100%) | 0 (0%) |
| Cefepime or Meropenem or PIP-TAZO + Azithromycin or Levofloxacin | 42 (66.6 %) | 10.3 (17-5) | 14.4 (27-5) | 24 (57%) | 18 (43%) |
| Cefepime or Meropenem or PIP-TAZO + Azithromycin or Levofloxacin + Vancomycin or Linezolid | 18 (28.6 %) | 10.2 (30-5) | 14.14 (24-10) | 5 (27.8%) | 13 (72.2%) |
| **Azithromycin + Hydroxychloroquine – no. (%)** |  |  |  |  |  |
| YES | 50 (79.4 %) | 10.06 (30-5) | 14.38 (24-5) | 23 (46%) | 27 (54%) |
| NO | 13 (20.6 %) | 11.15 (30-7) | 14.62 (27-10) | 9 (69.2%) | 4 (30.8%) |
| **Days need to wean from Oxygen Therapy – no. (%)** |  |  |  |  |  |
| 2-3 | 15 (23.8 %) | 7.4 (13-5) | 12.73 (20-5) | 15 (100%) | 0 (0%) |
| 4-5 | 13 (20.6 %) | 7.85 (12-5) | 14.54 (27-8) | 9 (69.2%) | 4 (30.8%) |
| 6-7 | 7 (11.1 %) | 8.57 (13-6) | 12.86 (17-11) | 2 (28.6%) | 5 (71.4%) |
| 8-10 | 15 (23.8 %) | 9.93 (14-6) | 15.6 (23-10) | 4 (26.7%) | 11 (73.3%) |
| 11-14 | 9 (14.3 %) | 13.89 (17-10) | 14.33 (24-8) | 1 (11.1%) | 8 (88.9%) |
| more than 14 days | 4 (6.3 %) | 25.25 (30-17) | 19 (24-14) | 1 (25%) | 3 (75%) |
| **Outcome – no. (%)** |  |  |  |  |  |
| No ICU +Discharge Home | 59 (93.6%) |  |  |  |  |
| Yes ICU +Discharge Home | 2 (3.2 %) |  |  |  |  |
| No ICU + Long Stay | 1 (1.6 %) |  |  |  |  |
| Yes ICU + Long Stay | 1 (1.6%) |  |  |  |  |
| Death | 0 |  |  |  |  |

<sup>a</sup> including 12 patients on non re-breathing mask

**Table (5).** Severe Covid 19 Pneumonia with hypoxemia and characteristics of Empirical Antibiotics and Corticosteroids treatment

| Covid-19 Pneumonia with Hypoxemia < 90 %<br>(All patients need more than 3L oxygen therapy to maintain oxygen saturation more than 92%) |  | Empirical Antibiotics |  |  | Days of Corticosteroids Treatment – no. (%) |  |  |
| --- | --- | --- | --- | --- | --- | --- | --- |
|  |  | Ceftriaxone + Azithromycin or Levofloxacin<br>No. (%) / 3 (4.8 %) | Cefepime or Meropenem or PIP-TAZO + Azithromycin or Levofloxacin<br>No. (%) / 42 (66.6 %) | Cefepime or Meropenem or PIP-TAZO + Azithromycin or Levofloxacin + Vancomycin or Linezolid<br>No. (%) / 18 (28.6 %) | 3 Days<br>No. (%) / 37 (58.7 %) | 5 Days<br>No. (%) / 16 (25.4 %) | More Than 5 Days<br>No. (%) / 10 (15.9 %) |
| **Age – no. (%)** |  |  |  |  |  |  |  |
| 18-40 | 15 (23.8 %) | 0 | 13 (86.7%) | 2 (13.3%) | 11 (73.3%) | 4 (26.7%) | 0 (0%) |
| 41-60 | 39 (61.9 %) | 3 (7.7%) | 23 (59%) | 13 (33.3%) | 22 (56.4%) | 10 (25.6%) | 7 (18%) |
| 60-80 | 9 (14.3 %) | 0 (0%) | 6 (66.7%) | 3 (33.3%) | 4 (44.5%) | 2 (22.2%) | 3 (33.3%) |
| **Sex – no. (%)** |  |  |  |  |  |  |  |
| Male | 55 (87.3 %) | 3 (5.5%) | 37 (67.3%) | 15 (27.2%) | 32 (58.2%) | 13 (23.6%) | 10 (18.2%) |
| Female | 8 (12.7 %) | 0 (0%) | 5 (62.5%) | 3 (37.5%) | 5 (62.5%) | 3 (37.5%) | 0 (0%) |
| **Comorbidity or Coexisting conditions – no. (%)** |  |  |  |  |  |  |  |
| NO | 26 (41.3 %) | 2 (7.7%) | 18 (69.2%) | 6 (23.1%) | 17 (65.4%) | 5 (19.2%) | 4 (15.4%) |
| YES | 37 (58.7 %) | 1 (2.7%) | 24 (64.9%) | 12 (32.4%) | 20 (54.1%) | 11 (29.7%) | 6 (16.2%) |
| **D-dimer Level – no. (%)** |  |  |  |  |  |  |  |
| < 1000 | 38 (60.3 %) | 3 | 24 | 11 | 20 | 12 | 6 |
| 1000 – 3000 | 21 (33.3 %) | 0 | 17 | 4 | 15 | 3 | 3 |
| > 3000 | 4 (6.3 %) | 0 | 1 | 3 | 2 | 1 | 1 |
| **Oxygen Therapy at Admission – no (%)** |  |  |  |  |  |  |  |
| Ambient Air | 0 |  |  |  |  |  |  |
| Low Flow Oxygen 3L oxygen or less | 0 |  |  |  |  |  |  |
| Low Flow Oxygen 4L – 9 L oxygen | 32 (50.8 %) | 3 (9.4%) | 24 (75%) | 5 (15.6%) | 26 (81.25%) | 6 (18.75%) | 0 (0%) |
| Low Flow Oxygen 10L oxygen or more | 31(49.2 %)<sup>a</sup> | 0 (0%) | 18 (58.1%) | 13 (41.9) | 11 (35.5%) | 10 (32.25%) | 10 (32.25%) |
| **Anti-inflammatory and Antiviral Treatment – no. (%)** |  |  |  |  |  |  |  |
| Anticoagulant | 0 |  |  |  |  |  |  |
| Anticoagulant + Hydroxychloroquine | 0 |  |  |  |  |  |  |
| Corticosteroids + Anticoagulant + Hydroxychloroquine | 59 (93.7 %) |  |  |  |  |  |  |
| Corticosteroids + Anticoagulant + Lopinavir | 4 (6.3 %) |  |  |  |  |  |  |
| **Days of Corticosteroids Treatment – no. (%)** |  |  |  |  |  |  |  |
| 3 days | 37 (58.7 %) | 3 (81.2%) | 28 (75.6%) | 6 (16.2) |  |  |  |
| 5 days | 16 (25.4 %) | 0 (0%) | 10 (62.5%) | 6 (37.5%) |  |  |  |
| More than 5 days | 10 (15.9 %) | 0 (0%) | 4 (40%) | 6 (60%) |  |  |  |
| **Empirical Antibiotics – no. (%)** |  |  |  |  |  |  |  |
| Ceftriaxone + Azithromycin or Levofloxacin | 3 (4.8 %) |  |  |  | 3 (100%) | 0 (0%) | 0 (0%) |
| Cefepime or Meropenem or PIP-TAZO + Azithromycin or Levofloxacin | 42 (66.6 %) |  |  |  | 28 (66.7%) | 10 (23.8%) | 4 (9.5%) |
| Cefepime or Meropenem or PIP-TAZO + Azithromycin or Levofloxacin + Vancomycin or Linezolid | 18 (28.6 %) |  |  |  | 6 (33.3%) | 6 (33.3%) | 6 (33.3%) |
| **Azithromycin + Hydroxychloroquine – no. (%)** |  |  |  |  |  |  |  |
| YES | 50 (79.4 %) | 3 (6%) | 33 (66%) | 14 (28%) | 28 (56%) | 13 (26%) | 9 (18%) |
| NO | 13 (20.6 %) | 0 (0%) | 9 (69.3%) | 4 (30.7%) | 9 (69.3) | 3 (23%) | 1 (7.7%) |
| **Days need to wean from Oxygen Therapy – no. (%)** |  |  |  |  |  |  |  |
| 2-3 | 15 (23.8 %) | 2 (33.3%) | 12 (80%) | 1 (6.7%) | 14 (93.3%) | 1 (6.7%) | 0 (0%) |
| 4-5 | 13 (20.6 %) | 0 (0%) | 11 (84.6%) | 2 (15.4%) | 10 (76.9%) | 3 (23.1%) | 0 (0%) |
| 6-7 | 7 (11.1 %) | 0 (0%) | 5 (71.4%) | 2 (28.6%) | 4 (57.1%) | 2 (28.6%) | 1 (14.3%) |
| 8-10 | 15 (23.8 %) | 1 (6.7%) | 8 (53.3%) | 6 (40%) | 8 (53.3%) | 6 (40%) | 1 (6.7%) |
| 11-14 | 9 (14.3 %) | 0 (0%) | 5 (55.6%) | 4 (44.4%) | 1 (11.1%) | 3 (33.3%) | 5 (55.6%) |
| more than 14 days | 4 (6.3 %) | 0 (0%) | 1 (25%) | 3 (75%) | 0 (0%) | 1 (25%) | 3 (75%) |
| **Outcome – no. (%)** |  |  |  |  |  |  |  |
| No ICU +Discharge Home | 59 (93.6 %) | 3 (5.1%) | 40 (67.8%) | 16 (27.1%) | 37 (62.7%) | 16 (27.1%) | 6 (10.2%) |
| Yes ICU +Discharge Home | 2 (3.2 %) | 0 (0%) | 1 (50%) | 1 (50%) | 0 (0%) | 0 (0%) | 2 (100%) |
| No ICU + Long Stay | 1 (1.6 %) | 0 (0%) | 0 (0%) | 1 (100%) | 0 (0%) | 0 (0%) | 1 (100%) |
| Yes ICU + Long Stay | 1 (1.6%) | 0 (0%) | 1 (100%) | 0 (0%) | 0 (0%) | 0 (0%) | 1 (100%) |
| Death | 0 (0%) |  |  |  |  |  |  |

<sup>a</sup> including 12 patients on non re-breathing mask

Moderate COVID-19 pneumonia group received Enoxaparin, antiviral drugs, empirical antibiotics for pneumonia, and standard treatment for comorbidity. Male patients were 50 (76.9 %) and female patients were 15 (23.1 %). A 34 (52.3 %) patients have comorbidity, 25 (38.5%) patients have Diabetes Mellitus and 2 (3.1 %) pregnant ladies. A 19 (29.2 %) patients were on low flow oxygen therapy, 3L oxygen or less to maintain oxygen saturation more than 92. 42 (64.6 %) patients received azithromycin, and hydroxychloroquine treatment and 23 (35.4 %) patients received Lopinavir/Ritonavir. The length of stay mean was 6.7 days (18 – 3), and virus clearance time mean was 14.3 days (24 – 10). All patients in the group discharged home with no major or minor bleeding complication or significant complication. All patients have 28 days follow up, or more after discharge without readmission or complication noticed.

The length of stay was higher among patients with comorbidity, and the mean was 7.08 (18-3) days. It also was higher among patients who received Lopinavir/Ritonavir only, and the mean was 7.59 (18-3) days, where the mean was 5.95 (14-3) days among patients received azithromycin and hydroxychloroquine treatment. Virus clearance time was prolonged among patients had low flow oxygen therapy with mean of 15.21 (24-10) day and prolonged among patients who received Lopinavir/Ritonavir only, and the mean was 15.45 (24-10) day, where the mean was 13.14 (24-10) day among patients received azithromycin and hydroxychloroquine treatment. Patients with comorbidity 13 (8.25%) needed more Low flow oxygen therapy than patients with no comorbidity.

Severe COVID-19 pneumonia group received methylprednisolone, Enoxaparin, antiviral drugs, empirical antibiotics for pneumonia, and standard treatment for comorbidity. Male patients were 55 (87.3 %) and female patients were 8 (12.7 %). 37 (58.7 %) patients have comorbidity, and 24 (38.1%) patients have Diabetes Mellitus. 32 (50.8 %) patients were on low flow oxygen therapy, 4-9L oxygen, and 31 (49.2 %) patients were on low flow oxygen therapy, 10L oxygen or more, including 12 patients on the non-rebreathing mask. Patients received methylprednisolone were 37 (58.7 %) for 3 days, 16 (25.4 %) for 5 days and 10 (15.9 %) for more than 5 days. 50 (79.4 %) patients received azithromycin, and hydroxychloroquine treatment and 13 (20.6 %) patients received Lopinavir/Ritonavir. Length of stay mean was 9.7 days (30 – 3), and virus clearance time mean was 14.3 (27 – 6). A 59 (93.6 %) patients in the group discharged home with no major or minor bleeding complication or septic shock. The most common complication was hyperglycemia among diabetic patients. There are two patients transferred to ICU due to respiratory failure and managed with non-invasive ventilation (BiPAP) for three days, then transferred back to the medical ward and discharged home. There is one patient in medical ward needs low flow oxygen therapy and methylprednisolone for more than 21 days due to unknown reason or confirmed significant lung disease or infection, but Bactrim was given empirically for possible pneumocystis pneumonia, then improved and discharged home. There is also one patient with diabetes mellitus and ischemic heart disease transferred to ICU due respiratory failure and managed with an invasive ventilator for 23 days and ended with a tracheostomy tube and severe lung fibrosis, currently he is conscious and pulmonary rehabilitation started. All patients have 28 days follow up, or more after discharge without readmission or complication noticed.

Length of stay was higher among patients with comorbidity, and the mean was 11.30 (30-6) day. It also was higher among patients who received 10L or more low flow oxygen therapy, and the mean was 12.47 (30-5) day. It also was high among patients who received Lopinavir/Ritonavir only, and the mean was 11.15 (30-7) day, where the mean was 10.06 (30-5) day among patients received azithromycin and hydroxychloroquine treatment. Virus clearance time was prolonged among patients who had low flow oxygen therapy with a mean of 15.03 (24-8) day and prolonged among patients who received methylprednisolone for more than five days, and the mean was 16.20 (24-8) day. Patients with comorbidity 22 (59.5%) needed more Low flow oxygen therapy, 10L oxygen, or more than patients with no comorbidity.

D-Dimer level was measured in all patients on admission in both groups—no significant association between D-Dimer level and hypoxemia, length of stay, or virus clearance time.

## Discussion

The clinical course of COVID-19 could be divided into three phases: viremia phase, acute phase (pneumonia phase) and severe or recovery phase.^7^

Patients with competent immune functions and without apparent risk factors (old age, comorbidities, etc.) may generate effective and adequate immune responses to suppress the virus in the first or second phase without immune over-reaction. In contrast, patients with immune dysfunction may have a higher risk of failing the initial phase and becoming severe or critical type with higher mortality.8

Therefore, treatment of COVID-19 should be based on the staging of patients, and the window of opportunity may lie between the first and the second phases, when clinical deterioration is observed with evidence of abrupt inflammation and hypercoagulable states. With no proved antivirals, early intervention has mainly focused on the correct timing of disease stages and implementing ways to stop or slow disease progression. Once the patients enter the critical status, nothing could be relied on other than comprehensive management.^8^

The combined use of anti-inflammatory and antiviral drugs may be more effective than using either modality alone. Based on in vitro evidence for inhibiting SARS-CoV-2 replication and blocking SARS-Co-2 infection-induced pro-inflammatory cytokine production.^9^

### Clinical-Therapeutic Staging Proposal ^10,11^

- Stage I: Mild (Early Infection) Administration of steroid during the early infection could increase viral replication and perhaps delay development of adaptive immunity. This might be expected to be detrimental.
- Stage IIa: Moderate (Pulmonary Involvement without Hypoxia)
- Stage IIb: Moderate (Pulmonary Involvement with Hypoxia) Low-dose steroid administration during the pulmonary involvement might be expected to be beneficial (by blunting the severity of inflammation and thereby preventing a severe hyper-inflammation phase).
- Stage III: Severe (Systemic Hyperinflammation)

For those patients who develop a marked hyper-inflammation phase, low-dose steroid might be inadequate to treat this. Higher doses of steroid or targeted immunosuppressive (e.g. tocilizumab) could be necessary to treat established hyper-inflammation. However, higher doses of steroids have greater side-effects – so delaying steroid administration until Stage III could result in missing the window of optimal intervention

Low peripheral capillary oxygen saturation (Spo2; with the cutoff of 90%) after receiving oxygen support along with the presence of dyspnea were found to be a strong predictor of mortality.^12^ Exertional dyspnea, an easily assessed symptom, is associated with death in patients with COVID-19 associated pneumonia independently of age and sex.^12^ Among patients who developed severe disease, the mean duration to develop dyspnea is 5 − 8 days, Acute respiratory distress syndrome (ARDS) from 8 –12 days, ICU admission is 10 –12 days. Some patients can rapidly deteriorate one week after illness onset. Mortality among patients admitted to the ICU is 39% –72%, and the Median length of hospitalization among survivors is 10 –13 days.^13,14^ Acute respiratory distress syndrome (ARDS) is the leading cause of mortality in COVID-19 pneumonia.^15^ Therefore, COVID-19 patients with exertional dyspnea with or without hypoxia should be admitted to hospitals, evaluated for pulmonary involvement clinically and radiologically, and treated early with anti-inflammatory and antiviral drugs.

Proper management of COVID-19 mandates a better understanding of disease pathogenesis. Two main features preceding severe respiratory failure associated with COVID-19: the first is macrophage activation syndrome-like state; the second is defective antigen-presentation driven by interleukin-6.^2^

The peculiar clinical course of CAP caused by SARS-CoV-2, including the sudden deterioration of the clinical condition 7– 8 days after the first symptoms, generates the hypothesis that this illness is driven by a unique pattern of immune dysfunction that is likely different from sepsis. The features of lymphopenia with hepatic dysfunction and increase of D-dimers in these patients with severe disease further support this hypothesis^.2,16^

It was reported that IL-6 and IL-8 could cause hypercoagulation, leading to scattered fibrin clots, shortening the clot dissolution time, and maximizing the dissolution rate.^17^ It was also observed that severe COVID-19 patients had higher levels of IL-6,^18^, suggesting that the hypercoagulation status of COVID-19 patients may be related to the elevated levels of cytokines. Pathological observations support the current concept of a hypercoagulative state in critically ill patients, showing that the frequency of pulmonary micro-thrombosis is high.^19^ Because organ dysfunction is mainly limited in lung and virus is the primary pathogen, the coagulation feature of severe COVID-19 might not be identical with sepsis in general.^20^

American Society of Hematology recommends LMWH should be considered in ALL patients (including non-critically ill) who require hospital admission for COVID-19 infection, in the absence of any contraindications like active bleeding, platelet count <25×109/L, monitoring advised in severe renal impairment and abnormal PT or aPTT is not a contraindication.^21^Despite the lack of quality published evidence, many institutional protocols have adopted an intermediate-intensity (i.e., administering the usual daily LMWH dose twice daily) or even a therapeutic-intensity dose strategy for thromboprophylaxis based on local experience.^22^

LMWH not only improves the coagulation dysfunction of COVID-19 patients but exerts an anti-inflammatory effect through reducing IL-6 and increasing lymphocyte%.^23^

If a MAS-like state exists and excessive IL-6 levels are detrimental, why shouldn’t corticosteroids be first-line therapy as these will vigorously suppress IL-6 and a raft of other cytokines? Although the recent open-label study from Wu and colleagues showed a benefit for corticosteroids,^24^ the consensus is that these should not be used based on clinical experience in SARS-CoV, MERS-CoV, and other infections including influenza and respiratory syncytial virus infection, where collectively there is evidence for delayed viral clearance.^25^ An early short course of methylprednisolone in patients with moderate to severe COVID-19 reduced escalation of care and improved clinical outcomes in a multi-center pre/post study evaluating the effect of a protocol involving early steroid administration within a health system in Michigan (including five hospitals).^26^

In this study, Enoxaparin was given as anti-inflammatory to COVID-19 pneumonia without major or minor bleeding complications. Enoxaparin dose based on D-Dimer level and oxygen saturation +/- methylprednisolone plus antiviral drugs is associated with better outcome in COVID-19 pneumonia. Enoxaparin was given for three to five days or more until oxygen saturation is more than 93%.

Methylprednisolone 40mg three or four times daily according to the severity of hypoxia. Patients oxygen saturation often improved to more than 93% on ambient air after 3 to 5 days of methylprednisolone treatment. During methylprednisolone treatment, oxygen saturation sometimes becomes abruptly normal, even if patients were on more than 5L oxygen therapy. Hyperglycemia, hypernatremia, and hypokalemia were monitored and treated. Routine sepsis monitoring, including bloodwork and monitoring for recurrence of inflammation and signs of adrenal insufficiency after stopping methylprednisolone, were performed. Some patients had high blood pressure, gastritis/duodenitis, and insomnia during methylprednisolone treatment. The rate of occurrence of infectious complications increases, and mild infection may become severe and fatal. Corticosteroids may also mask some signs of current infection. Latent infections may be activated, or there may be an exacerbation of intercurrent infections due to pathogens, including those caused by Amoeba, Candida, Cryptococcus, Mycobacterium, Nocardia, Pneumocystis, Toxoplasma, and fungal infection. There is one patient who was given methylprednisolone for more than 21 days, after ten days of steroids therapy, workup for possible infections were done, no confirmed significant lung disease or infection was found, and Bactrim was given empirically for possible pneumocystis pneumonia, then improved and discharged home.

Some patients were treated by methylprednisolone for ten days and more due to slowly improved hypoxemia. Those patients routinely monitored for pulmonary and extrapulmonary infections.

Antiviral drugs available at study time were hydroxychloroquine and Lopinavir/Ritonavir. Lopinavir/Ritonavir was given to patients who cannot take hydroxychloroquine because of prolonged QTc interval, old age, and multiple comorbidities, which explained the prolonged duration of virus clearance time and the length of stay in hospital inpatients taking Lopinavir/Ritonavir. The antiviral drugs with evidence of improving COVID-19 pneumonia can be combined with anti-inflammatory drugs to treat COVID-19 patients in hospitals.

Empirical antibiotics with standard community-acquired pneumonia regimen (ceftriaxone plus azithromycin or levofloxacin) in moderate COVID-19 pneumonia was given to 48 (73.8 %) patients. Most patients with diabetes mellitus (especially longstanding and uncontrolled) not improved on standard regimen or worse after 48 hours of admission, so hospital-acquired pneumonia regimen (Cefepime or Meropenem or PIP-TAZO + Azithromycin or Levofloxacin) was given and patients improved and discharged home. Extra days of length of stay, and virus clearance time, higher oxygen therapy among patients with comorbidity and moderate COVID-19 pneumonia. MRSA coverage was given to patients with a high risk of MRSA and not improved with hospital-acquired pneumonia regimen.

Diabetes is a risk factor for hospitalization and mortality of the COVID-19 infection. Diabetes was a comorbidity in 22% of 32 non-survivors in a study of 52 intensive care patients.^13^ In another study of 173 patients with severe disease, 16.2% had diabetes, and in further study of 140 hospitalized patients, 12% had diabetes.^27,28^ When comparing intensive care and non-intensive care patients with COVID-19, there appears to be a twofold increase in the incidence of patients in intensive care having diabetes.30 Mortality seems to be about threefold higher in people with diabetes compared with the general mortality of COVID-19 in China.^13,29^

In patients with severe COVID-19 pneumonia, empirical antibiotics with hospital-acquired pneumonia regimen (Cefepime or Meropenem or PIP-TAZO + Azithromycin or Levofloxacin) was given because of steroids therapy. Standard community-acquired pneumonia regimen with steroid therapy was failed many times because the patients have bilateral pulmonary involvement with hypoxemia. Comorbidity in severe COVID-19 pneumonia required more oxygen therapy and length of stay. MRSA coverage was given to patients with a high risk of MRSA and not improved with hospital-acquired pneumonia regimen. Empirical Bactrim was given once due to the possibility of pneumocystis pneumonia.

## CONCLUSION AND SUMMARY

Larger randomized studies and more analysis of patient subtypes to determine benefit from the use of combined anti-inflammatory (corticosteroids and Enoxaparin) and antiviral drugs treatment. It is hoped that the above analysis can provide a testable theoretical framework to allow for advances in our understanding and control of this deadly viral epidemic.

In conclusion, early use of a combined anti-inflammatory (corticosteroids and Enoxaparin) and antiviral drugs treatment, an inexpensive and readily available agents, in patients with moderate to severe COVID-19 pneumonia prevent complications of the disease and improve clinical outcomes. The combination therapy of anti-inflammatory and antiviral drugs can over great benefits to COVID-19 patients with ongoing COVID-19 pandemic and ICU bed and mechanical ventilator shortages.

## Data Availability

no

